# A 17-month longitudinal environmental sampling study carried out on public transport vehicles operating in England during the COVID-19 pandemic identified low levels of SARS-CoV-2 RNA contamination

**DOI:** 10.1101/2023.12.20.23300219

**Authors:** Paz Aranega-Bou, Thomas Pottage, Abigail Fenwick, Wilhemina D’Costa, Natalie F. Brown, Nicola Yaxley, Marco-Felipe King, Simon T. Parker, Daniel Miller, Martín López-García, Catherine J. Noakes, Ginny Moore, Allan Bennett

## Abstract

**Aims:** To monitor severe acute respiratory syndrome coronavirus-2 (SARS-CoV-2) RNA contamination in vehicles operating in England during the pandemic, to better understand transmission risk of SARS-CoV-2 on public transport.

**Methods and Results:** We collected 1,314 surface samples between December 2020 and April 2022 on trains and buses managed by five different transport operators. The presence of SARS-CoV-2 RNA was investigated through reverse transcription polymerase chain reaction (RT-PCR). SARS-CoV-2 RNA was found on 197 (15%) of the 1,314 surfaces sampled, including seat head rests, handholds, and air extract grilles, but the levels of RNA recovered on those samples (median value of 23.4, inter-quartile range: 14.3-35.4, *N* gene copies per extraction) made the presence of infectious virus at the time of sampling extremely unlikely. However, detection rates varied over time with peaks broadly coinciding with times of high community transmission, suggesting that people infected with SARS-CoV-2 when travelling on public transport could create opportunities for transmission.

**Conclusion:** During the pandemic, and as in other public spaces, low levels of SARS-CoV-2 RNA were found on surfaces associated with public transport.

**Impact statement:** The results of this study will inform modelling approaches and the implementation of mitigation strategies to minimise the risk of transmission of respiratory viruses in public transport.

## 1. Introduction

Public transport is an essential service, and it is associated with improved individual employment possibilities, a more active lifestyle and reduced air pollution (Saif et al., 2019). In 2019, public transport use in England equated to 33 rail journeys and 50 bus trips per person. These were taken for a variety of reasons including shopping, education, healthcare and leisure with over half of all rail journeys being for commuting and business purposes (Department for Transport, 2020; Lucas et al., 2019). In 2021, 22% of English households did not have access to a private car (Department for Transport, 2022) and therefore relied on public transport, walking or cycling for their transport needs (Lucas et al., 2019). Lowest income households are less likely to own a car, and it has been shown that inequalities in transport provision can stop people from accessing key local services and activities and contribute to social exclusion (Lucas et al., 2019). Due to the crucial nature of public transport, during the COVID-19 pandemic, it was essential to understand the potential transmission risk of SARS-CoV-2 and the effectiveness of mitigation strategies used in this setting.

As with other indoor settings where individuals can come into close contact with one another, use of public transport carries a risk of COVID-19 transmission (Gartland et al., 2022). Modelling work suggests that passengers in a subway carriage could be exposed to SARS-CoV-2 through the airborne route (via small aerosols) without having to be within 2 meters of the infectious source, the close-range route (via droplets or aerosols) or through exposure to contaminated fomites (Miller et al., 2022). The same study also identified potential mitigation strategies, including policies to prevent infectious passengers from travelling and/or to promote social distancing, the wearing of face coverings and frequent handwashing (Miller et al., 2022).

Data collection can inform policy decisions and refine modelling assumptions. Several studies have investigated the presence of SARS-CoV-2 RNA in public transport vehicles (Caggiano et al., 2021; Di Carlo et al., 2020; Hadei et al., 2021; Hoffman et al., 2022; Moreno et al., 2021). However, these studies have been carried out over short sampling timeframes. Longitudinal data collected over longer periods reflecting different situations (prevalence of infection, implemented mitigations, patronage, vaccination status in the population etc.) has been lacking. Here we report the results from a longitudinal sampling study in which 1,314 samples were collected from buses and trains operating in England between December 2020 and April 2022.

## 2. Methods

### 2.1. Transport operators

Transport operators, selected to give a spread of location and type of route, were approached to take part in the study. Two train operators (T1 and T2) and three bus operators (B1-B3) agreed to participate. Together these provided some geographical spread, operating long distance and local routes in areas of the Northeast, Northwest, South and Southwest of England. Vehicles sampled included passenger trains and both single-decker and double-decker buses.

All transport operators were implementing enhanced cleaning protocols during the sampling period which included daily cleaning of touch points and periodic heavy cleaning (approximately monthly). A range of products were used including BioHygiene sanitiser (Biological Preparations), Autoglym potassium hydroxide solution (Autoglym), Virabact solution (Cleenol), Selgiene Ultra (Selden), OdorBac Tec4 (OdorBac)and X-Mist (X-Mist Limited). Antimicrobial coatings were utilised by three operators, with both train operators (T1 and T2) applying Zoono Z71 Microbe Shield (Zoono) to contact surfaces on a monthly basis and operator B3 applying Zonitise Antimicrobial Spray (Zonitise) annually as per manufacturers’ instructions.

### 2.2. Sampling protocol

Samples were collected by operatives working for the transport operators following instructions provided by the study team at UKHSA. Sampling materials were prepared by the UKHSA team and delivered, via courier, to each of the five transport operators once a month. Details of this process can be found in the supplementary material.

Operatives were asked to take samples on a pre-specified date once a month when the vehicle had returned to depot after use at the end of the day and before it was subjected to daily terminal cleaning. Samples were collected using sterile cellulose sampling sponges with neutralising buffer (Technical Service Consultants Ltd.). On each sampling occasion, the internal surfaces of two buses or two second class carriages of the same train (designated as vehicle/carriage A and B) were sampled. Ten pre-determined samples were collected from each train carriage (Table 1) and eight from each bus (Table 2). All surfaces were in areas that could be occupied by passengers. None were associated with the driver’s cab. In each vehicle, multiple surface sites of the same type (e.g. handholds) were numbered. Those that were sampled were selected via a random number generator. For example, for operator B1, each bus had 16 handholds with a bell push and each month, two were selected at random to be sampled. Tables 1 and 2 provide the sampling coverage for each surface type on each vehicle/carriage. Whilst it was not possible to ensure the same vehicle was sampled each month, the vehicles that were sampled all covered the same or similar routes in each region.

**Table 1.**
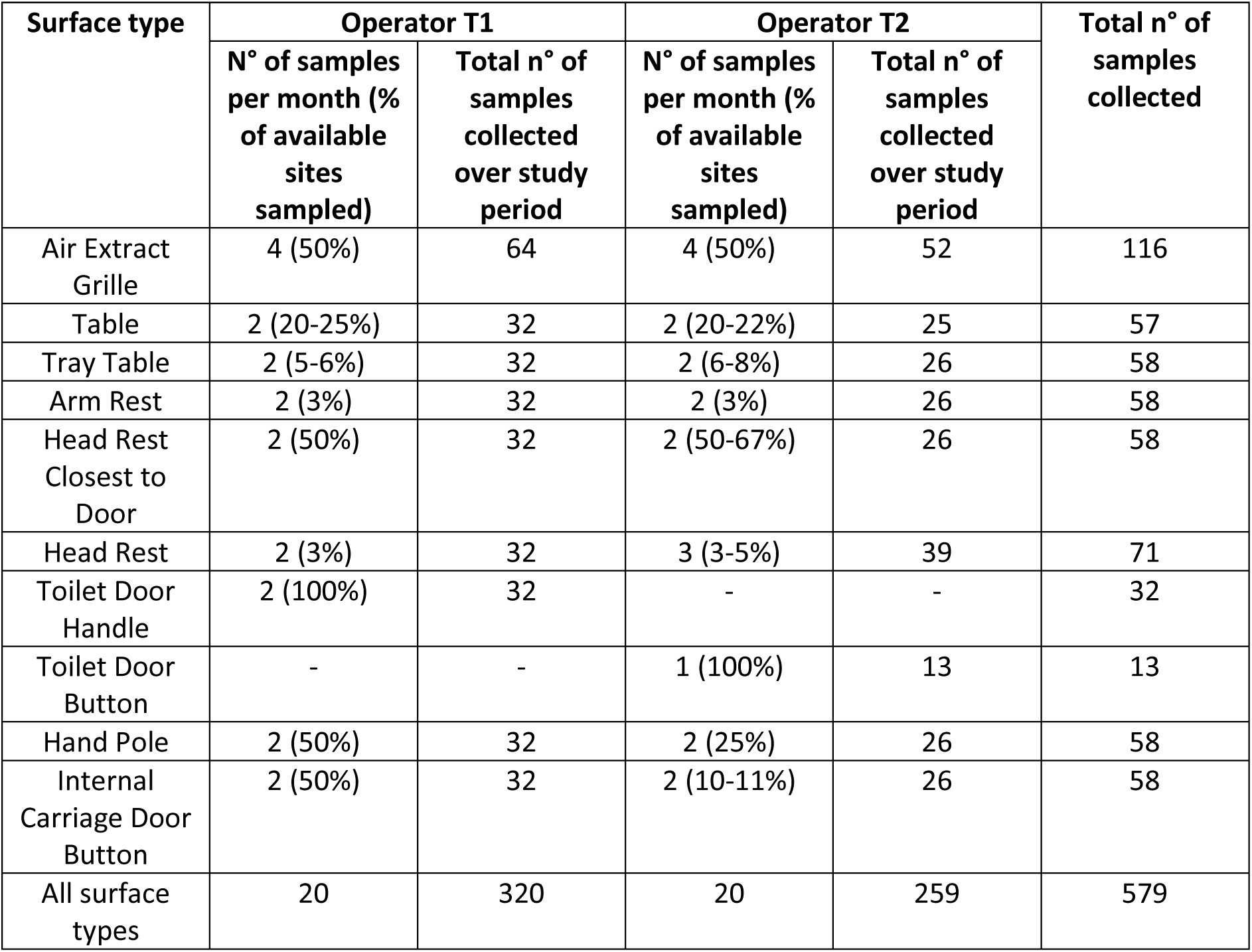
Overview of the different surface types sampled on passenger train carriages.

| Surface type | Operator T1 |  | Operator T2 |  | Total n° of samples collected |
| --- | --- | --- | --- | --- | --- |
|  | N° of samples per month (% of available sites sampled) | Total n° of samples collected over study period | N° of samples per month (% of available sites sampled) | Total n° of samples collected over study period |  |
| Air Extract Grille | 4 (50%) | 64 | 4 (50%) | 52 | 116 |
| Table | 2 (20-25%) | 32 | 2 (20-22%) | 25 | 57 |
| Tray Table | 2 (5-6%) | 32 | 2 (6-8%) | 26 | 58 |
| Arm Rest | 2 (3%) | 32 | 2 (3%) | 26 | 58 |
| Head Rest Closest to Door | 2 (50%) | 32 | 2 (50-67%) | 26 | 58 |
| Head Rest | 2 (3%) | 32 | 3 (3-5%) | 39 | 71 |
| Toilet Door Handle | 2 (100%) | 32 | - | - | 32 |
| Toilet Door Button | - | - | 1 (100%) | 13 | 13 |
| Hand Pole | 2 (50%) | 32 | 2 (25%) | 26 | 58 |
| Internal Carriage Door Button | 2 (50%) | 32 | 2 (10-11%) | 26 | 58 |
| All surface types | 20 | 320 | 20 | 259 | 579 |

**Table 2.** Overview of the different surface types sampled on buses.

| Surface type | Operator B1 |  | Operator B2 |  | Operator B3 |  | Total n° of samples collected |
| --- | --- | --- | --- | --- | --- | --- | --- |
|  | N° of samples per month (% of available sites sampled) | Total n° of samples collected over study period | N° of samples per month (% of available sites sampled) | Total n° of samples collected over study period | N° of samples per month (% of available sites sampled) | Total n° of samples collected |  |
| Head Rest | 6 (11%) | 96 | 4 (3%) | 60 | 4 (4%) | 60 | 216 |
| Handhold | 2 (33%) | 32 | 4 (11%) | 60 | 2 (10%) | 30 | 122 |
| Handhold High | 2 (14%) | 32 | - | - | - | - | 32 |
| Handhold Stairs | - | - | 2 (14%) | 30 | 2 (20%) | 30 | 60 |
| Handhold and Bell Push | 4 (13%) | 64 | 4 (14%) | 60 | 4 (13%) | 60 | 184 |
| Window Hopper | 2 (13%) | 32 | 2 (10%) | 30 | 2 (10%) | 29 | 91 |
| Table | - | - | - | - | 2 (25%) | 30 | 30 |
| All surface types | 16 | 256 | 16 | 240 | 16 | 239 | 735 |

Sampling sponges were returned to UKHSA via courier and were in transit at room temperature for between twelve hours and seven days before being received.

### 2.3. Sample processing

Once received, samples were stored at 4 °C or, if not processed within 24 h of receipt, at −80 °C. Sample sponges were retained within the associated bag and massaged by hand for ∼15 seconds. Approximately 1.5 mL of the neutralising buffer associated with each sponge was transferred to a 2-mL screw cap tube using a 2-mL stripette. A 100 µL and a 10 µL aliquot was inoculated on Columbia Blood Agar plates (CBA; E&O Laboratories Ltd.) and incubated at 37 °C for 48 hours. After incubation, bacteria were enumerated.

RNA was extracted using the QIAamp viral RNA mini kit (Qiagen) following the manufacturer’s instructions. Unused sampling sponges were extracted and processed as negative controls. Duplicate aliquots were screened for detection of SARS-CoV-2 RNA by RT-PCR using the VIASURE SARS-CoV-2 Real Time PCR Detection kit (CerTest Biotec) which detects the *N* and *ORF1ab* genes and includes an internal control, a positive control and a negative control to monitor inhibition in samples and for quality assurance. Samples were quantified using a standard curve on the *N* target, run on each plate. A sample was considered positive when amplification of at least one target (Ct<40) was detected in both replicates and weak positive when amplification (of at least one target) was detected in a single replicate. RNA extracts associated with ‘weak positive’ samples were re-analysed and considered positive if amplification was detected in both replicates. Samples were also re-analysed if the standard deviation for the internal control Ct values for the two replicates was >0.5. The mean quantification value for all aliquots with RNA detection is reported.

Selected samples with the highest *N* gene copies per extraction were subjected to whole genome sequencing. Sequencing was carried out by the UKHSA Pathogen Genomics Service at Porton Down, following the ARTIC network protocol (https://www.protocols.io/view/ncov-2019-sequencing-protocol-v3-locost-bp2l6n26rgqe/v3).

### 2.4. Statistical analysis

Firstly, a chi-square test of independence was done to check for a relationship between vehicle type (trains versus buses) and the proportion of positive samples. Logistic regression models were then used to further investigate the relationship between the likelihood of finding a positive sample and the variables vehicle type and surface type as not all surfaces were shared between buses and trains. Both vehicle type and surface type were included as predictor variables. This allowed the assessment of the effect of each variable while controlling for the effect of the other. All statistical tests were two-sided and the significance level was set at 0.05.

## 3. Results

### 3.1. SARS-CoV-2 RNA was detected on train and bus surfaces

Between December 2020 and April 2022, 1,314 surface samples were collected from trains belonging to 2 operators (T1-T2) and buses belonging to 3 different operators (B1-B3). In total, 92 buses and 58 train carriages were sampled. All samples were analysed using RT-PCR. Overall, SARS-CoV-2 RNA was detected on 197 (15%) surfaces. The percentage of samples collected from trains that contained SARS-CoV-2 RNA was the same regardless of operator (19%). The percentage of samples collected from buses that contained SARS-CoV-2 RNA was 16%, 15% and 3% for operators B1, B2 and B3 respectively (Figure 1a). Upon controlling for sample location (i.e. surface type), the type of vehicle (train or bus) did not significantly influence the odds of obtaining a positive SARS-CoV-2 RNA result, as determined by logistic regression analysis (p=0.423). This suggests that the likelihood of finding positive samples does not significantly differ between trains and buses when taking into account the specific location of the sample within the vehicle.

**Fig. 1.**
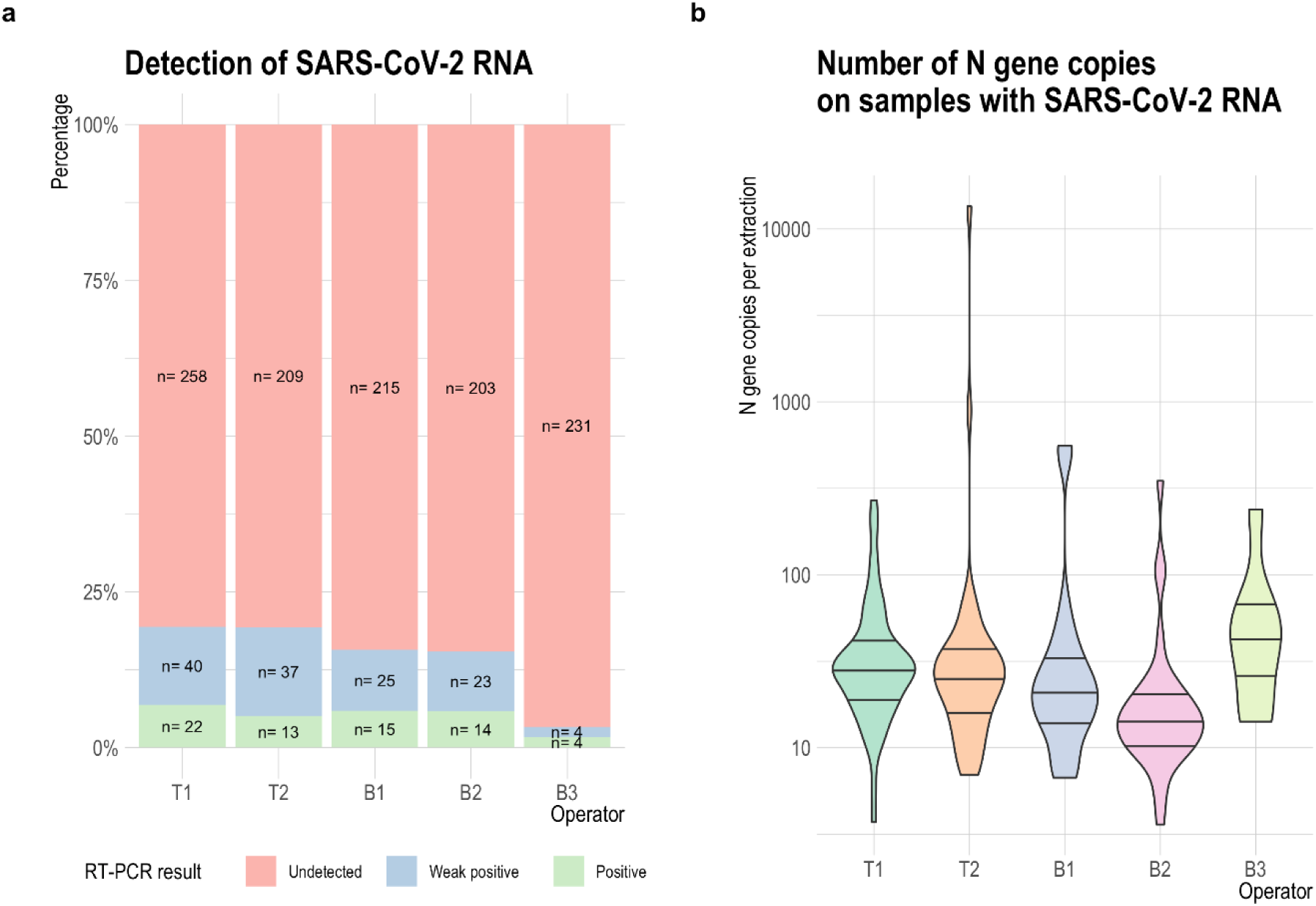
Detection of SARS-CoV-2 RNA in samples collected on train carriages (T1-T2) and buses (B1-B3) in England between December 2020 and April 2022. (a) Percentage of samples where SARS-CoV-2 RNA was detected in duplicate aliquots (positive) or in single aliquots (weak positive). (b) Number of *N* gene copies per extraction detected in samples containing SARS-CoV-2 RNA. The lines indicate the median and the interquartile range.

On surfaces where SARS-CoV-2 RNA was detected, the median number of *N* gene copies per extraction ranged from 14.3 (operator B2; n=240) to 42.4 (operator B3; n=239). The mean ranged from 29.2 (operator B2) to 338.5 (operator T2; n=259). While there were a few outliers, the concentration of SARS-CoV-2 RNA detected in 93% of positive samples equated to less than 100 *N* gene copies per extraction (Figure 1b). The highest concentrations of RNA were detected on a tray table (13,504 *N* gene copies per extraction) and an arm rest (892 *N* gene copies per extraction) sampled by operator T2 in March 2022 and a head rest (558 *N* gene copies per extraction) sampled by operator B1 in August 2021. These three samples were subjected to whole genome sequencing but none yielded enough reads to provide a complete genome sequence. All surfaces were contaminated with high numbers of bacteria (87% of which were contaminated at levels >40,000 CFU/surface).

### 3.2. SARS-CoV-2 RNA detection varied over time

Detection of SARS-CoV-2 RNA varied over time, regardless of operator and vehicle type. Timepoints associated with the highest positivity rates (i.e. percentage of samples contaminated with SARS-CoV-2 RNA) included January 2021 (47%; when only operators T1 and B1 were collecting samples), March 2022 (31%) and January 2022 (26%) (Figure 2; Supplementary figure 1). There was no correlation between the number of passengers that had travelled on the buses sampled and the percentage of surfaces contaminated with SARS-CoV-2 RNA (Supplementary figure 2). Train passenger data were not available. However, there was some indication that surface positivity rate increased with increasing numbers of people testing positive for COVID-19 within the community (Office for National Statistics positivity rate). This was particularly true for operator T2 (Supplementary figure 3).

**Fig. 2.**
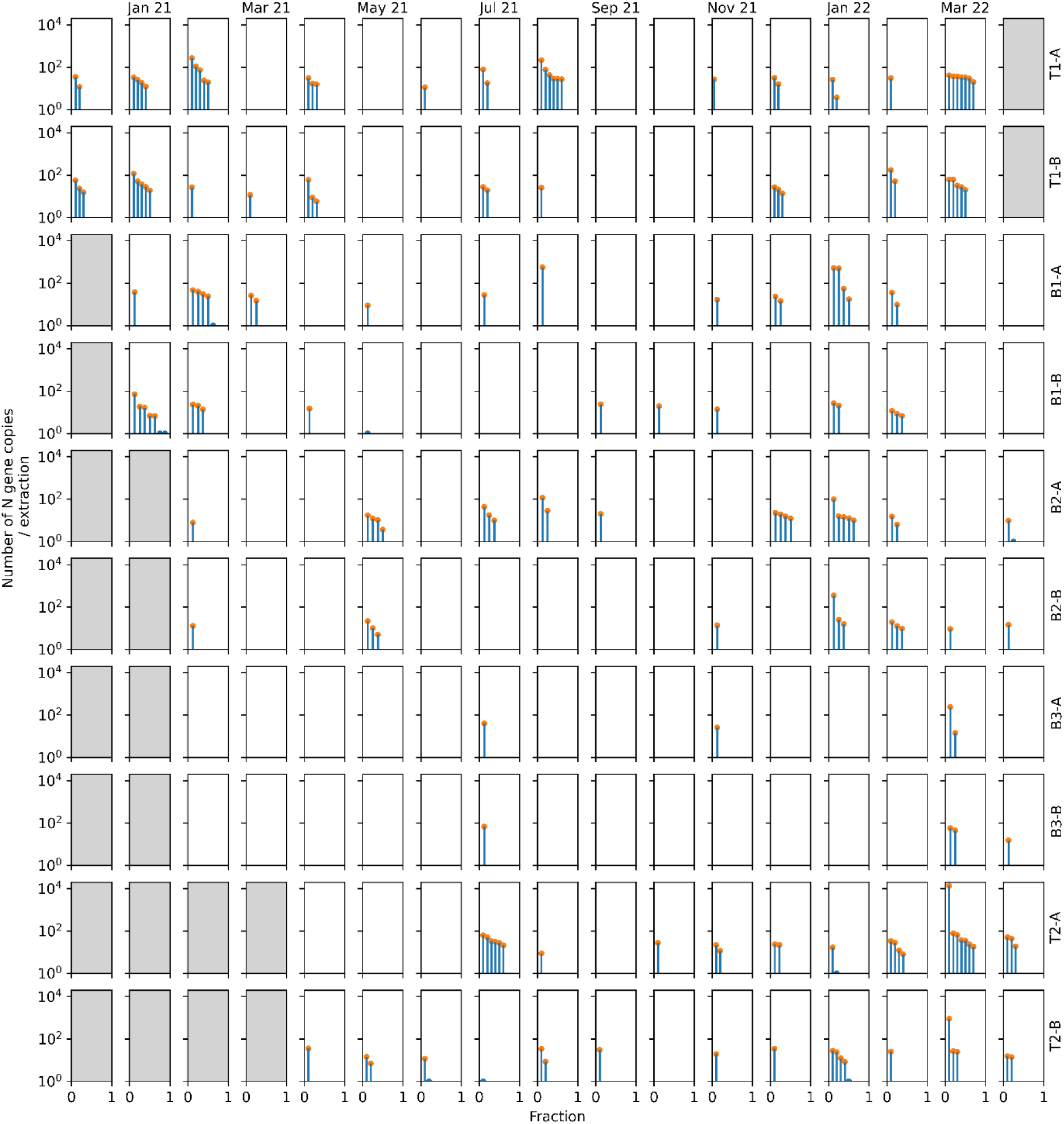
Detection and quantification (number of *N* gene copies per extraction) of SARS-CoV-2 RNA on the surfaces of train carriages (T1-T2) and buses (B1-B3) operating in England between December 2020 and April 2022. Each transport operator sampled two vehicles on each sampling occasion. Each row represents a vehicle, and each rectangle displays the results for one month, with each sample containing SARS-CoV-2 RNA represented on the x-axes (blue column) and the number of *N* gene copies per extraction for each of the samples represented on the y-axes (orange circle). Samples where SARS-CoV-2 RNA was detected but could not be quantified are represented with a blue semi-circle on the x-axis (e.g B1-B on May 2021). When no data was available for a vehicle on a certain month the square is shaded. Note that it was not possible to sample the same vehicles each month.

Widespread contamination, defined as detection of SARS-CoV-2 RNA in at least 50% of the samples collected on an individual vehicle, was observed in 5 (5%) of the 92 buses and 8 (14%) of the 58 train carriages sampled. In January 2021, 7 (88%) of the 8 surfaces sampled in vehicle B1-B were contaminated with SARS-CoV-2 RNA. Likewise, in March 2022, 7 (70%) of the 10 surfaces sampled in both carriage T1-A and carriage T2-A were contaminated with SARS-CoV-2 RNA. Conversely, SARS-CoV-2 RNA was not detected in any sample collected from 51 (55%) of the 92 buses sampled or from 15 (26%) of the 58 train carriages (Figure 2).

### 3.3. Some types of surface were more likely to be contaminated with SARS-CoV-2 RNA than others

SARS-CoV-2 RNA was detected on all sampled surface types at least once over the course of the study (Table 3). On trains, 129 seat head rests were sampled. Fifty-eight of these were associated with seats located close to the internal carriage door whilst 71 were located elsewhere within the carriage. SARS-CoV-2 was more likely to be detected on those closest to the door (27% vs 8%). Similarly, sample positivity was higher for shared tables (26%) than for tray tables (17%) and for toilet door handles (16%) than for toilet door buttons (8%).

**Table 3.** Detection of SARS-CoV-2 RNA on the different surface types sampled on buses and trains in England between December 2020 and April 2022. Sampling locations are ordered by most to least detection for each transport operator.

| VEHICLE TYPE | SURFACE TYPE | OPERATOR | N° SAMPLES COLLECTED | N° SAMPLES WHERE SARS-COV-2 RNA WAS DETECTED | % SAMPLES WHERE SARS-COV-2 RNA WAS DETECTED |
| --- | --- | --- | --- | --- | --- |
| TRAIN CARRIAGE | Head Rest Closest to Door | T1, T2 | 58 | 16 | 27% |
|  | Table | T1, T2 | 57 | 15 | 26% |
|  | Air Extract Grille | T1, T2 | 116 | 28 | 24% |
|  | Arm Rest | T1, T2 | 58 | 14 | 24% |
|  | Internal Carriage Door Button | T1, T2 | 58 | 10 | 17% |
|  | Tray Table | T1, T2 | 58 | 10 | 17% |
|  | Toilet Door Handle | T1 | 32 | 5 | 16% |
|  | Hand Pole | T1, T2 | 58 | 7 | 12% |
|  | Head Rest | T1, T2 | 71 | 6 | 8% |
|  | Toilet Door Button | T2 | 13 | 1 | 8% |
| BUS | Handhold | B1, B2, B3 | 122 | 21 | 17% |
|  | Handhold High | B1 | 32 | 4 | 13% |
|  | Head Rest | B1, B2, B3 | 216 | 27 | 13% |
|  | Handhold and Bell push | B1, B2, B3 | 184 | 21 | 11% |
|  | Window Hopper | B1, B2, B3 | 91 | 8 | 9% |
|  | Handhold Stairs | B2, B3 | 60 | 3 | 5% |
|  | Table | B3 | 30 | 1 | 3% |

Only 3% of the internal carriage door button surfaces sampled by operator T1 were found to be contaminated with SARS-CoV-2 RNA, compared to 35% of those sampled by operator T2 (supplementary table 1). This might have been due to vehicles operated by operator T2 having a higher number of internal doors therefore increasing opportunities for interaction with the open buttons. Air extract grilles were also frequently contaminated with SARS-CoV-2 RNA, particularly in vehicles operated by operator T1 (31% for operator T1 vs. 15% for operator T2) (supplementary table 1). On buses, those surfaces that could be used as a handhold were the most contaminated (Table 3).

Whilst differences existed, our overall test did not find a significant association with sample positivity (Likelihood ratio chi-squared = 20.075, df = 14, p = 0.1278). However, individual surface types did show significant associations. Specifically, samples taken from ‘Handhold and Bell Push’ (Estimate = −1.015, p = 0.0458), ‘Handhold Stairs’ (Estimate = −1.693, p = 0.0237), ‘Head Rest’ (Estimate = −1.028, p = 0.0142), and ‘Window Hopper’ (Estimate = −1.333, p = 0.0231) were less likely to be positive compared to the ‘Air Extract Grille’.

Despite the lack of overall statistical significance, these individual associations suggest that the likelihood of SARS-CoV-2 RNA can vary between specific surface types. As such, while our analysis did not find surface type as a whole to be a significant predictor of sample positivity, specific surface types do appear to be associated with the likelihood of finding the virus.

## 4. Discussion

Here we describe the results of a longitudinal sampling study conducted on train carriages and buses operating in England between December 2020 and April 2022. SARS-CoV-2 RNA was detected in 84 (58%) of the 146 vehicles sampled and, although viability was not assessed, the low levels of SARS-CoV-2 RNA detected suggest that no live virus was present on any of the surfaces at the time of sampling. However, it is acknowledged that RNA degradation may have occurred over time and it is possible that infectious virus may have been present on surfaces sometime prior to sampling. The inability to successfully sequence samples containing comparatively high concentrations of RNA confirmed the potential for viral genetic material to partially degrade. Moreover, when being returned to the laboratory, samples were usually in transit at room temperature for between 12 and 36 hours but occasionally for up to seven days, thereby increasing the potential for some RNA degradation. In previous studies, infectious SARS-CoV-2 virus has been isolated from surfaces in close proximity to individuals who have tested positive for COVID-19 in the hospital (Ahn et al., 2020), the domestic setting (Marcenac et al., 2021) and during an experimental infection study (Zhou et al., 2023). In a study investigating household transmission, infectious SARS-CoV-2 virus was also isolated from the hands of a contact who subsequently tested positive one day later (Derqui et al., 2023). While laboratory studies have reported that SARS-CoV-2 can survive on surfaces and skin for hours to days when left undisturbed (Hirose et al., 2020; Paton et al., 2021; Riddell et al., 2020), the persistence of infectious virus under more realistic conditions remains to be fully investigated. In contrast, there is evidence to suggest that SARS-CoV-2 RNA can persist on a wide range of porous and nonporous surfaces for long periods. Salido et al., (2021) used heat inactivated viral particles to monitor the persistence of SARS-CoV-2 RNA over a period of one week. No decay of SARS-CoV-2 RNA was observed on many commonly used materials. Paton et al., (2021) contaminated surfaces with live SARS-CoV-2 virus and reported a 90% (one log_10_) reduction in SARS-CoV-2 RNA concentration after three weeks, despite viability being lost within hours or days. A study that involved the sampling of 17 farms did not detect any SARS-CoV-2 RNA in 246 swabs collected from animals or in 93 swabs collected from farm workers but traces of SARS-CoV-2 RNA were detected in 11 of 77 concurrent environmental samples collected from stable walls, barkers and milking parlours, suggesting that surface contamination from previous infection events had persisted (Cardinale et al., 2022). In a home where one adult and one child had been isolating with COVID-19, 46% of 24 surfaces were contaminated with SARS-CoV-2 RNA one month after symptom resolution (Maestre et al., 2021). Therefore, in the current study, the detection of residual RNA may not reflect a recent contamination event.

Understanding how different settings and activities contribute to transmission of SARS-CoV-2 is key to better targeted public health interventions and recommendations. Contact tracing has determined that households are responsible for most transmission but there is limited data for other social settings, including workplaces and schools (Thompson et al., 2021). Linking transmission events to a particular journey in public transport is particularly challenging due to the random and dynamic group of people using multiple vehicles for variable amounts of time, often to travel to other social settings (Gartland et al., 2022). Nonetheless, outbreaks of SARS-CoV-2 have been linked to bus travel in China (Luo et al., 2020a; Shen et al., 2020) and a study carried out in Norway reported an association between use of public transport and SARS-CoV-2 infection (Ellingjord-Dale et al., 2022). On the other hand, other studies have reported little to no transmission. Studies tracing public transport contacts in China cited attack rates of 0.1% to 2.1% compared to 4.2% for private cars (Liu et al., 2020; Luo et al., 2020b). A school in USA observed no transmission on 15 buses transporting 462 students on a school route, even though 37 students, 1 driver and 1 aide were determined to have travelled while infectious at some point (Ramirez et al., 2021). In these instances, the implementation of mitigation strategies (e.g. use of face coverings, ventilation and sanitation) may have reduced the risk of transmission (Liu et al., 2020; Luo et al., 2020b; Ramirez et al., 2021).

In our study, all transport operators applied enhanced cleaning protocols during the sampling period. Moreover, three operators supplemented routine cleaning with antimicrobial coatings applied to high-touch surfaces. Operator T1 and T2 applied Zoono Z71 Microbe Shield surface sanitizer monthly and operator B3 applied Zonitise antimicrobial spray yearly, as recommended by the manufacturers. Despite this, the number of bacteria recovered from all surfaces, regardless of presence or type of coating was consistently high (87% of all samples were contaminated at levels >40,000 CFU/surface) suggesting limited effectiveness of antimicrobial coatings in this setting. While samples were taken before daily terminal cleaning, high bacterial levels demonstrate the ease with which passengers contaminate the environment. We previously showed that when Zoono was used to coat a range of materials relevant to public transport, the deposition of organic debris severely reduced its virucidal efficacy against ɸ6 (a surrogate for SARS-CoV-2) (Aranega-Bou et al., 2023). As part of the same study, we found that Zonitise, when applied to stainless steel or polystyrene test surfaces, was ineffective against ɸ6 even in the absence of organic debris (previously unpublished data, supplementary table 2). However, its effect on RNA was not assessed, and whilst many factors could account for the less frequent contamination of vehicles operated by operator B3, degradation of RNA cannot be ruled out. Further research into appropriate mitigation strategies for public transport is warranted to ensure the safety of this crucial service, including the development of cleaning guidelines and standards to facilitate effective cleaning in this setting.

Previous sampling studies have described higher recovery of SARS-CoV-2 RNA on surfaces in close prolonged contact with people infected with SARS-CoV-2 in households (Marcenac et al., 2021) and hospitals (Moore et al., 2021). In our study, some surface types were contaminated with SARS-CoV-2 RNA more frequently than others, although these differences were not found to be statistically significant when considering the type of surface as a whole. In general, those surfaces found to be contaminated most often were those associated with handholds (i.e., surfaces that are gripped) and/or where there is opportunity for prolonged contact (e.g., tables and arm rests) or frequent contact (e.g., head rest of seats close to the carriage door, which passengers are often seen holding when waiting to leave the train). These surface types might facilitate the dispersal of SARS-CoV-2 in this environment via hand contact and self-inoculation could occur if facial mucosas are touched after contact with a contaminated surface. A previous study investigating surface touch networks and surface contamination with a tracer bacterium in an office observed high contamination on the office door handle and chair seatbacks, both surfaces that are gripped during contact, and determined that both surfaces contributed to contamination spread, alongside hand-to-hand contact (Wang et al., 2021). In an observational study on the subway of Mexico City, 89% of the 120 passengers selected at random for observation contacted a pole during their trip, with a mean of 4.4 contacts per passenger every 10 minutes (Vargas-Robles et al., 2020). Both studies also reported individuals touching their facial mucosas, with 17% of subway passengers doing so during their trip with a mean of 0.2 contacts per passenger every 10 minutes (Vargas-Robles et al., 2020; Wang et al., 2021). Future work linking behavioural observations in public transport to microbiological sampling data might elucidate the contribution of the surface types identified in this study to contamination spread and provide some context to allow more informative interpretation of results.

Studies with volunteers have shown that SARS-CoV-2 RNA can be shed in aerosol particles in exhaled breath by people with a SARS-CoV-2 infection during respiratory activities such as breathing and talking, particularly during early stages of infection (Adenaiye et al., 2022; Ma et al., 2021; Tan et al., 2023). Previous studies in public transport have detected SARS-CoV-2 RNA in air samples taken on buses and from associated air conditioning filters (Hadei et al., 2021; Hoffman et al., 2022; Moreno et al., 2021). Others have not (Di Carlo et al., 2020), probably reflecting differences in sampling methodologies, prevalence of COVID-19 in the community, mitigation strategies (e.g. use of face coverings) and/or passenger behaviour. Swabbing transportation air extract grilles could be a good alternative to indirectly sample the air, as SARS-CoV-2 RNA has previously been found on the surfaces of ventilation grilles and other air handling unit components in various locations housing COVID-19 patients (Maestre et al., 2021; Mouchtouri et al., 2020; Santarpia et al., 2020). In this study, 31% and 15% of air extract grilles sampled by operators T1 and T2 respectively were contaminated with SARS-CoV-2 RNA, suggesting that viral RNA and potentially viable SARS-CoV-2 virus had been present in the air before being captured on the surface of the air extract over time. Air extract grilles were more likely to be contaminated than other surfaces. However, this most likely reflects less frequent cleaning in comparison to high touch surface sites, and the accumulation of contamination over time.

The percentage of samples with SARS-CoV-2 RNA was very similar for both train operators (19%) and for operators B1 and B2 (16% and 15% respectively) but a lower rate was reported for operator B3 (3%). The reasons for the lower detection rate for operator B3 are unknown. Previous studies carried out on trains and buses over shorter timeframes (1 day to 11 days) reported detection rates of between 0% and 43% (Caggiano et al., 2021; Cardinale et al., 2022; Di Carlo et al., 2020; Moreno et al., 2021). Whilst these studies included comparatively fewer samples (ranging from 30 to 150) they also illustrate that variable levels of contamination can be detected in different vehicles, geographic locations and pandemic waves. In our study, detection of SARS-CoV-2 RNA varied over time with peaks mostly coinciding with times of high community transmission. Detection was highest during the winter months of 2020/2021 (when the Alpha (B.1.1.7) variant emerged in England) and in the winter and spring of 2021/2022 (concurrent to the spread of the Omicron (B.1.1.529) variant in England). A smaller peak was also observed in summer 2021 which correlates with the spread of the Delta (B.1.617.2) variant (Wellcome Sanger Institute). It is likely that the number of individuals travelling on public transport whilst infected with COVID-19 will increase during times of high community transmission. Similarly, Zhang et al., (2022) observed a correlation between the number of COVID-19 cases at a university and the presence of SARS-CoV-2 RNA in air and surface samples collected in the campus gym, bus, lab office and classroom. Differences in the prevalent variant over the sampling period might have also affected the detection rates. It has been suggested that infection with the Alpha and Omicron variants might be associated with increased shedding of SARS-CoV-2 RNA, although the results are confounded by high variability between individuals and the low number of individuals included in the studies (Adenaiye et al., 2022; Tan et al., 2023).

The main limitation of this study is the scarcity of metadata in relation to the parameters that might have affected the recovery of SARS-CoV-2 RNA from surfaces including the number and location of passengers, their infection status and demographic characteristics, behaviour including the use of face coverings and vaccination status, cleaning frequency and the ambient temperature and relative humidity of the vehicles, which can affect viral persistence (Biryukov et al., 2020). However, the number of daily passengers on buses was available and no correlation was found with the detection rate, suggesting that other parameters had a bigger influence. We also did not assess the competency of those carrying out the sampling; if the sampling instructions were followed appropriately or their sampling technique, which might have introduced bias (Hedman et al., 2020). To simplify the sampling and for ease of use by the samplers, we chose cellulose sponges to facilitate the sampling of large areas and operators were instructed to sample each site in its entirety (e.g., the whole length of a handhold). However, our sampling method made RNA quantification difficult, and results are presented as copies per extraction instead of copies per surface area. Another limitation for using this data in modelling is that the sampling efficiency for the different materials that were sampled is unknown and likely to vary (Hardison et al., 2023; Jansson et al., 2020). Sampling efficiency is predicted to have a large effect on the amount of SARS-CoV-2 RNA detected on surfaces (Supplementary figure 4) but it is very challenging to determine experimentally due to the high number of variables involved.

In conclusion, while we detected SARS-CoV-2 RNA on surfaces at levels which were not indicative of the presence of live virus, the occasional widespread contamination on some public transport vehicles suggests that people who travel on public transport when infected with SARS-CoV-2 may contaminate the environment and create opportunities for onward transmission. Appropriate mitigation strategies should be identified and implemented, particularly during times of high community transmission. However, we found no evidence of public transport contamination with SARS-CoV-2 RNA being higher in comparison to other public spaces including university campuses, public squares, business premises and bus stations (Abrahão et al., 2021; Cardinale et al., 2022; Harvey et al., 2021; Mihajlovski et al., 2022; Zhang et al., 2022).

## Author contributions

Conceptualization: PAB, TP, CN, GM and AB; Data curation: PAB; Formal analysis and Visualization: PAB, MFK, SP, DM, MLG; Funding acquisition: TP, CN, GM, AB; Investigation: PAB, AF, WDC, NB, NY; Methodology: PAB, TP, NY, GM; Project administration: PAB, TP, CN, GM, AB; Writing-original draft: PAB; Writing-review and editing: all authors.

## Acknowledgements

We would like to thank all the transport operators that participated in the study and, in particular, the operatives that collected samples over the 17-month sampling period. We would also like to thank the “UKHSA Pathogen Genomics Service – Porton Down” for carrying out the sequencing work. The views expressed in this article are those of the authors and are not necessarily those of UKHSA or of the Department of Health and Social Care.

## Data availability

The data underlying this article will be shared on reasonable request to the corresponding author.

