## Supplemental material for "A 17-month longitudinal environmental sampling study carried out on public transport vehicles operating in England during the COVID-19 pandemic identified low levels of SARS-CoV-2 RNA contamination"

**Supplementary material**

**Methods**

**Sampling protocol**

Instructions to sampling operators were provided during a face-to-face session on the type of vehicle to be sampled or, when COVID-19 restrictions prevented the study team from traveling, an online session via Microsoft Teams. When necessary (e.g. change of operator staff) instructions were cascaded within the transport operator company. Regardless of the format, all appropriate sampling materials were provided for use during the session to allow familiarisation by the operatives. All materials were packaged within a cardboard courier box and included: prelabelled sterile sampling sponges with neutralising buffer (Technical Service Consultants Ltd.), disinfectant wipes (PDI Sani-Cloth 70% Alcohol wipes), nitrile gloves, sampling sheets detailing where samples should be taken for each vehicle, floorplan of the vehicles to be sampled annotated with the sample sites to be sampled, sampling methodology picture sheet (supplementary appendix A), sealable bags (DGP Pathoseal 95) for the packaging of the sampling sponges compliant to UN3373 regulations, a waste bag and a return label. Each session included a discussion describing (i) the contents of the box; (ii) the information required by the study team (e.g. route sampled); (iii) how to use the sampling sheet and vehicle maps to locate the designated sampling sites and (iv) how to carry out the sampling.

Operators were instructed to put on gloves and sample each site using a single sampling sponge. After returning the sponge to its bag, operators were instructed to wipe the surface that had been sampled, the sampling sponge bag and their gloves with a disinfectant wipe before moving on to the next sampling site (supplementary appendix A). Whilst operatives had the opportunity to follow the sampling procedure and to ask any questions prior to starting the study, there was no follow-up. The UKHSA team did not assess whether sampling was undertaken according to the instructions provided.

**Results**

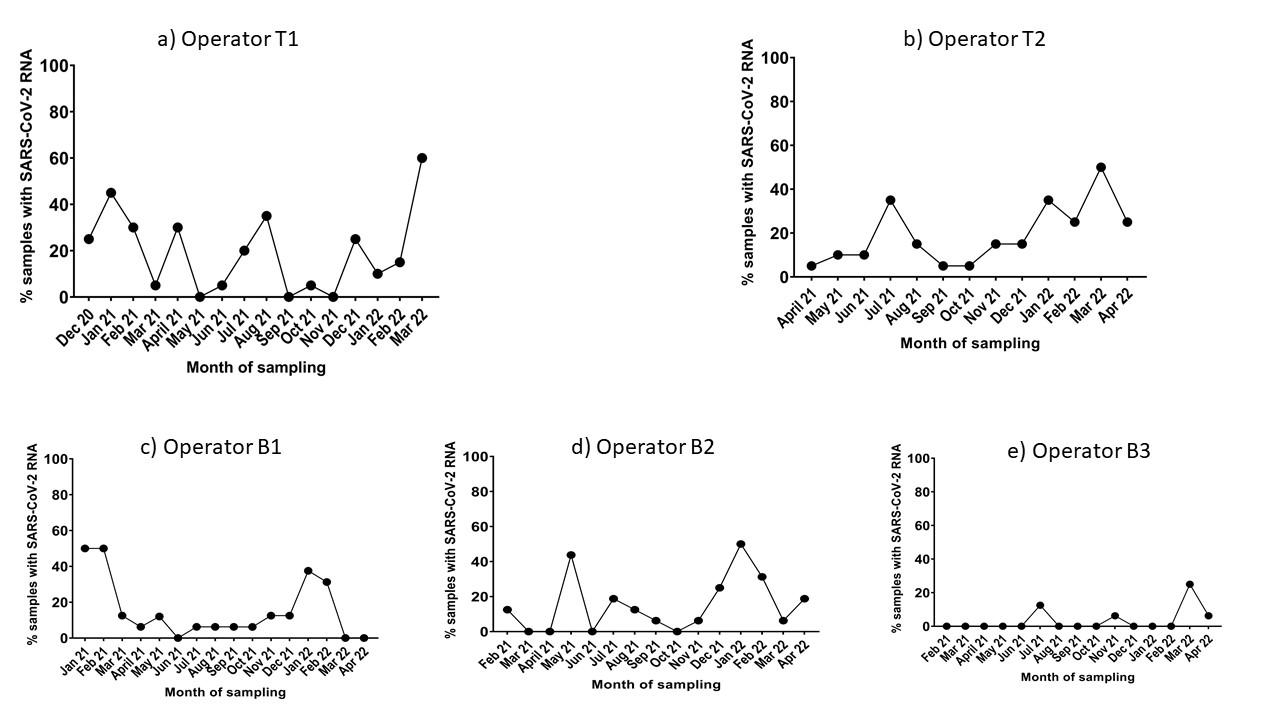

**Supplementary figure 1.** Percentage of monthly samples collected by each train operator (T1-T2) and bus operator (B1-B3) where SARS-CoV-2 RNA was detected. The number of sampling episodes completed by each operator was 16 (14/12/2020, 19/01/2021, 16/02/2021, 16/03/2021, 19/04/2021, 18/05/2021, 22/06/2021, 20/07/2021, 24/08/2021, 23/9/2021, 02/11/2021 (reported as October 2021), 22/11/2021, 21/12/2021, 24/01/20212, 03/03/2022 (reported as February 2022) and 29/03/2022) for operator T1, 13 (12/04/2021, 12/05/2021, 07/06/2021, 13/07/2021, 10/08/2021, 12/09/2021, 11/10/2021, 08/11/2021, 13/12/2021, 10/01/2022, 15/02/2022, 21/03/2022 and 11/04/2022) for operator T2, 16 (04/01/2021, 02/02/2021, 02/03/2021, 06/04/2021, 10/05/2021, 07/06/2021, 05/07/2021, 02/08/2021, 07/09/2021, 12/10/2021, 01/11/2021, 06/12/2021, 11/01/2022, 07/02/2022, 09/03/2022 and 05/04/2022) for operator B1, 15 (02/02/2021, 02/03/2021, 06/04/2021, 04/05/2021, 08/06/2021, 06/07/2021, 03/08/2021, 07/09/2021, 14/10/2021, 02/11/2021, 09/12/2021, 12/01/2022, 08/02/2022, 08/03/2022 and 05/04/2022) for operator B2 and 15 (10/02/2021, 11/03/2021, 14/04/2021, 16/05/2021, 17/06/2021, 15/07/2021, 18/08/2021, 14/09/2021, 13/10/2021, 11/11/2021, 08/12/2021, 11/01/2022, 17/02/2022, 17/03/2022 and 11/04/2022) for operator B3.

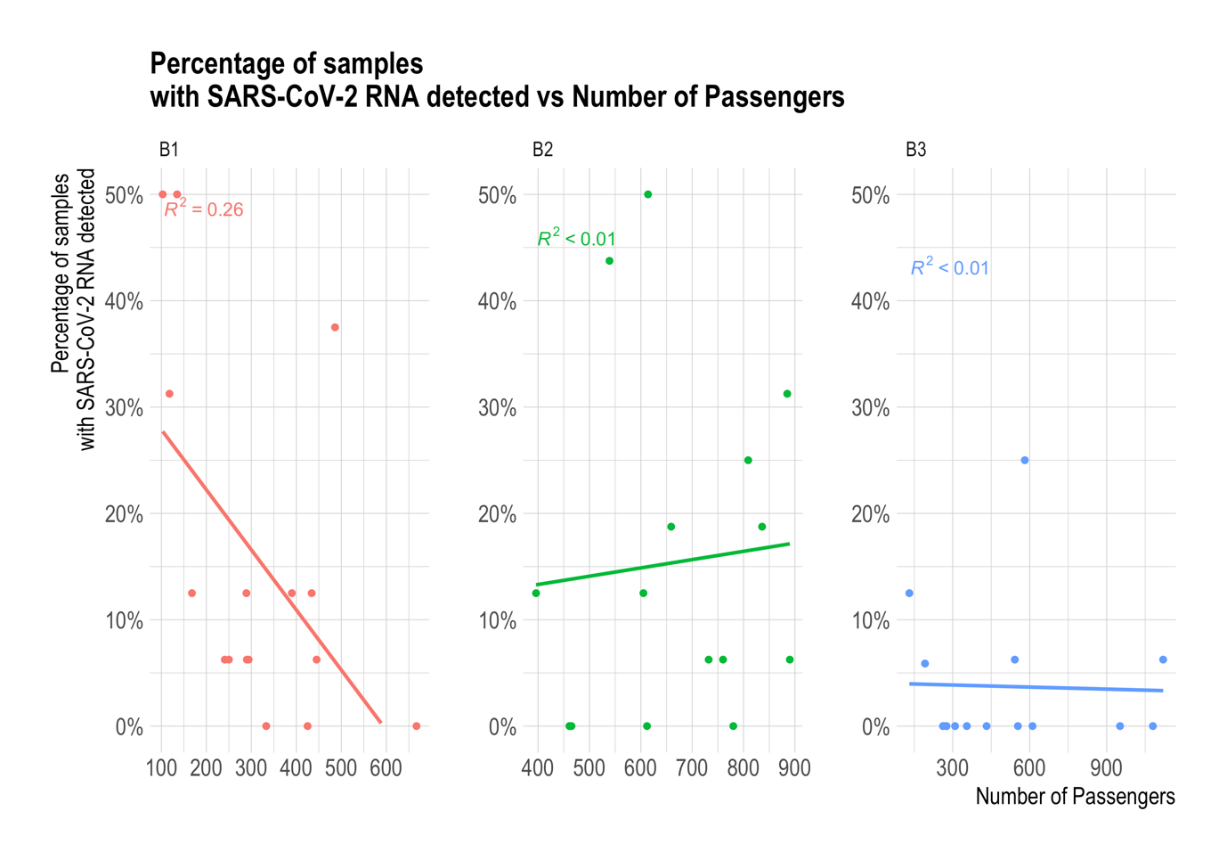

**Supplementary figure 2.** Percentage of samples with SARS-CoV-2 RNA detected vs. number of passengers travelling on the buses sampled on the day the sample were collected.

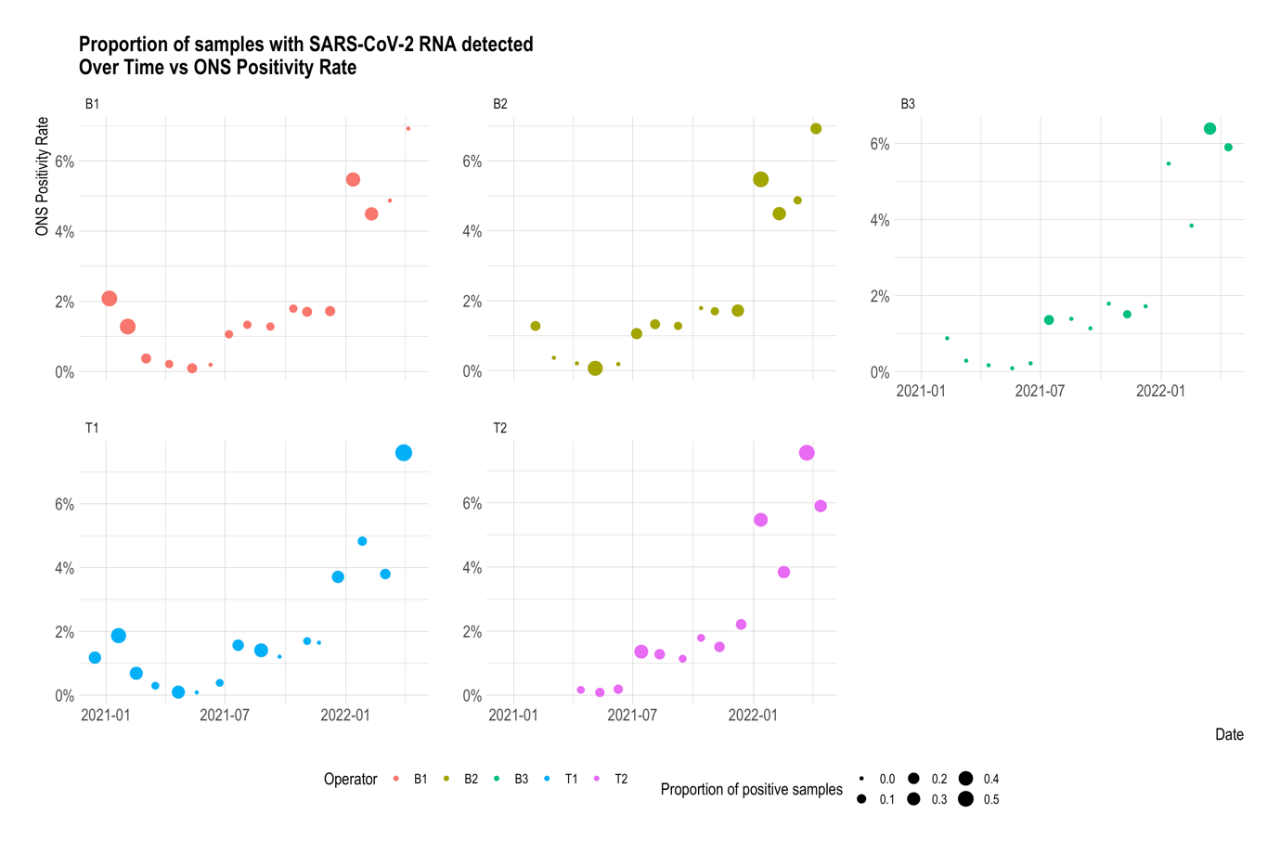

**Supplementary figure 3.** Proportion of samples with SARS-CoV-2 detected over time vs. ONS positivity rate.

**Supplementary table 1:** Detection of SARS-CoV-2 RNA on the different surface types sampled by two train operators (T1-T2) and three bus operators (B1-B3) in England between December 2020 and April 2022. Sampling locations are ordered by most to least detection for each transport operator.

| Operator | Sampling location | N° samples collected | N° samples where SARS-CoV-2 RNA was detected | % samples where SARS-CoV-2 RNA was detected |
| --- | --- | --- | --- | --- |
| T1 | Air Extract Grille | 64 | 20 | 31% |
|  | Head Rest Closest to Door | 32 | 8 | 25% |
|  | Table | 32 | 7 | 22% |
|  | Tray Table | 32 | 7 | 22% |
|  | Arm Rest | 32 | 6 | 19% |
|  | Hand Pole | 32 | 5 | 16% |
|  | Toilet Door Handle | 32 | 5 | 16% |
|  | Head Rest | 32 | 3 | 9% |
|  | Internal Carriage Door Button | 32 | 1 | 3% |
| T2 | Internal Carriage Door Button | 26 | 9 | 35% |
|  | Table | 25 | 8 | 32% |
|  | Arm Rest | 26 | 8 | 31% |
|  | Head Rest Closest to Door | 26 | 8 | 31% |
|  | Air Extract Grille | 52 | 8 | 15% |
|  | Tray Table | 26 | 3 | 12% |
|  | Hand Pole | 26 | 2 | 8% |
|  | Head Rest | 39 | 3 | 8% |
|  | Toilet Door Button | 13 | 1 | 8% |
| B1 | Handhold | 32 | 7 | 22% |
|  | Handhold and Bell Push | 64 | 12 | 19% |
|  | Head rest | 96 | 15 | 16% |
|  | Handhold High | 32 | 4 | 13% |
|  | Window Hopper | 32 | 2 | 6% |
| B2 | Handhold | 60 | 12 | 20% |
|  | Window Hopper | 30 | 5 | 17% |
|  | Head Rest | 60 | 9 | 15% |
|  | Handhold and Bell Push | 60 | 8 | 13% |
|  | Handhold Stairs | 30 | 3 | 10% |
| B3 | Handhold | 30 | 2 | 7% |
|  | Head Rest | 60 | 3 | 5% |
|  | Table | 30 | 1 | 3% |
|  | Window Hopper | 29 | 1 | 3% |
|  | Handhold and Bell Push | 60 | 1 | 2% |
|  | Handhold Stairs | 30 | 0 | 0% |

**Supplementary table 2:** Efficacy of Zonitise against ɸ6 when applied to stainless steel or polystyrene test surfaces by spraying 24 hours prior to ɸ6 inoculation. Efficacy is expressed as log10 reduction calculated by subtracting the mean ɸ6 PFU recovered on coated test surfaces from the mean ɸ6 PFU recovered from non-coated test surfaces after a 120-minute contact time (n=3 coupons per condition).

| Stainless steel | Polystyrene |
| --- | --- |
| 0.1 | 0.0 |

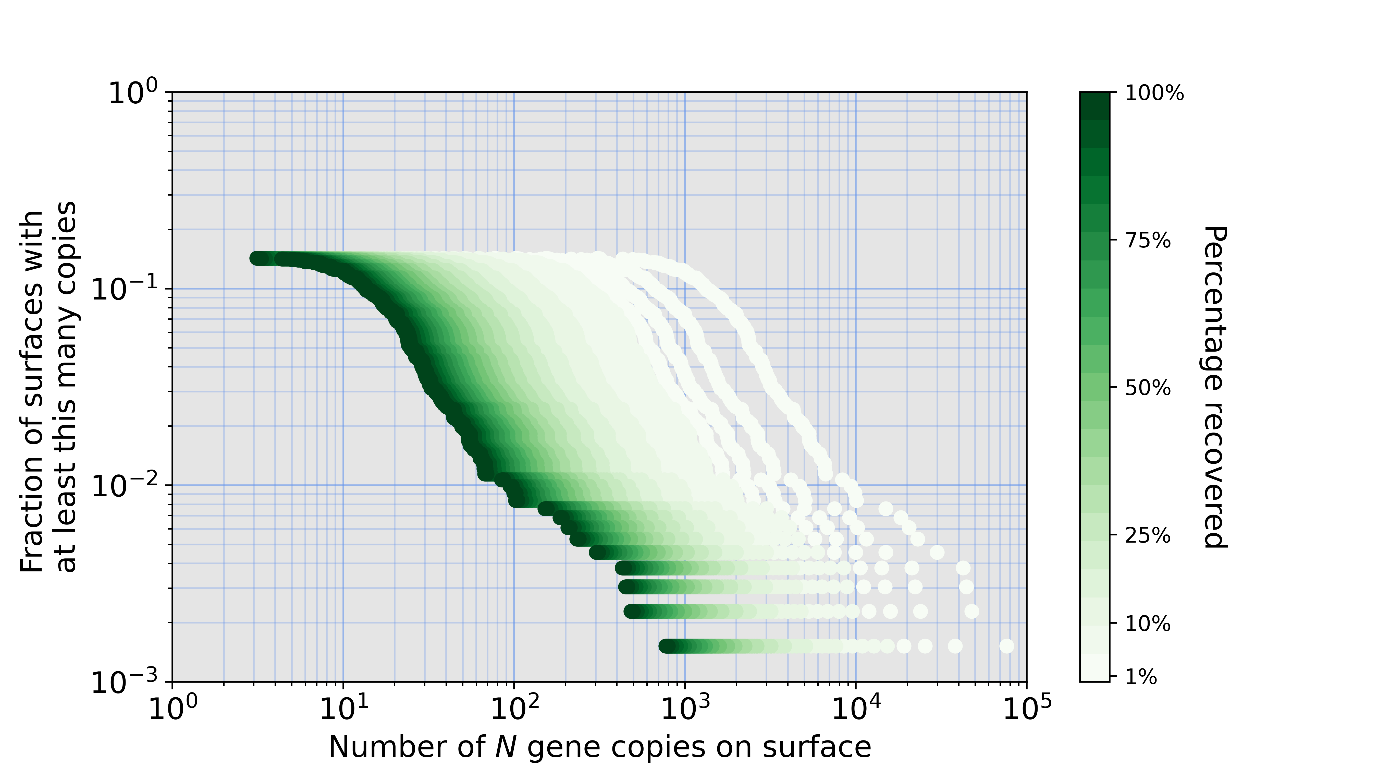

**Supplementary figure 4:** Change in the fraction of surfaces with at least as many gene copies as the value on the horizontal with the percentage of RNA recovered from the surface during sampling indicated by the colour. It is observed that reasonable estimates for the recovery percentages (1-100%) result in a significant difference in the estimated surface contamination.

**Supplementary appendix A**

**TRACK PROJECT - Protocol for the sampling of transport vehicles**

Prior to sampling, the equipment required will be sent to your office:

- Sampling sheets and drawings – detailing where samples should be taken
- Prelabelled environmental sponges – one for each sampling site
- Nitrile gloves
- Disinfectant wipes
- Bag for the packaging of the used environmental sponges
- Waste bag
- Label to be attached to box to send samples to Porton Down

Not included: pen to record on sampling sheet and tape to attach label to box

| 1. Put on clean nitrile gloves  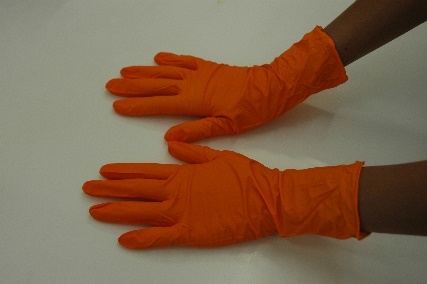 | 2. Check the sampling sheet to identify the environmental sponge to use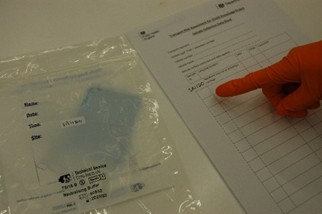 |
| --- | --- |
| 3. Tear open the sponge’s bag  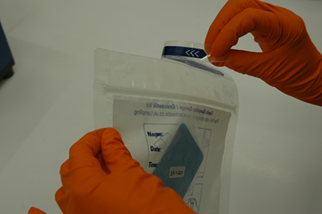 | 4. Remove the sponge from the bag  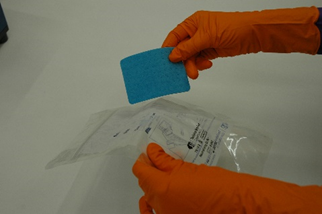 |
| 5. Wipe the sampling site with the sponge, using horizontal and vertical strokes, for larger areas use both side of the sponge  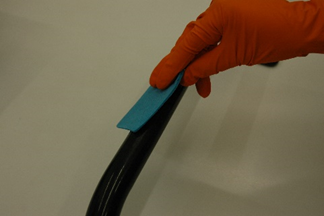 | 6. Seal sponge in bag using the zip lock. Make sure bag is intact and sealed  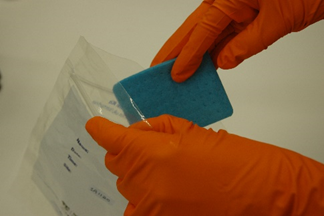 |
| 7. Wipe gloves with a disinfectant wipe  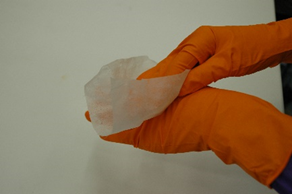 | 8. Wipe the exterior of the sponge bag with a disinfectant wipe  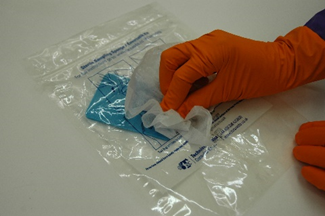 |
| 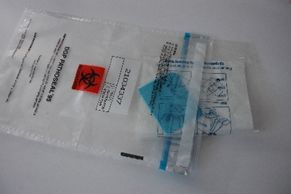9. Put the sponge sample into the larger collection bag | 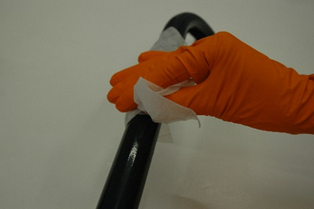10. Decontaminate the area sampled using the disinfectant wipes, ensure the area sampled is wetted with the disinfectant from the wipes. More than one wipe may be required |
| 11. Date and sign the sampling sheet to record the sample has been taken.  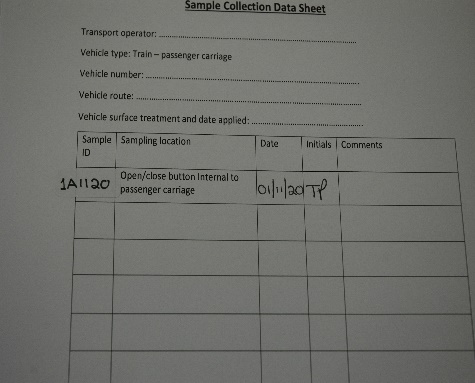 | 12. If necessary, write any comments in the box provided  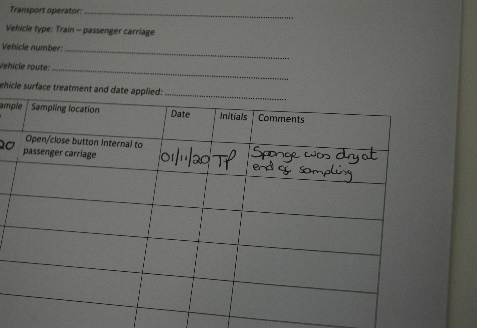 |
| 13. Disinfectant wipes can be placed in existing waste streams | 14. Repeat process from steps 3 to 13 for remaining sampling sites |
| 15. After all samples have been taken close collection bag with collected samples in and place in box for courier  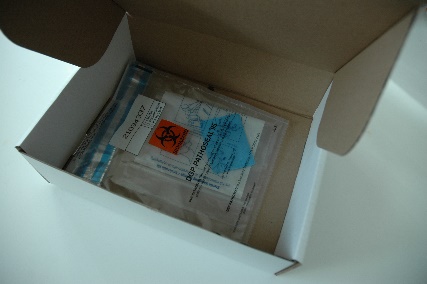 | Any questions please contact:  Thomas Pottage  |
